# Vaccine, Booster and Natural Antibody Binding to SARS-CoV-2 Omicron (BA.1) Spike Protein and Vaccine Efficacy

**DOI:** 10.1101/2022.07.12.22277539

**Authors:** Philip H. James-Pemberton, Mark W. Helliwell, Rouslan V. Olkhov, Shivali Kohli, Aaron C. Westlake, Benjamin M. Farrar, Ben J. Sutton, Nicholas D. Ager, Andrew M. Shaw

## Abstract

The SARS-CoV-2 Omicron variant (BA.1) has 25 unique mutations to the Spike glycoprotein, suggesting the efficacy of current vaccines against the new variant may be seriously degraded. A fully quantitative antibody binding study was performed for Spike Omicron (SO) and original Spike (S) proteins simultaneously on three cohorts of patients: convalescent following RT-PCR-confirmed infection in early 2020, double-vaccinated at ≥2 weeks, and vaccine boosters. The average (mode) of the booster cohort response distributions were 15.1 mg/L and 13.4 mg/L for S and SO, respectively, compared with the significantly lower double-vaccinated average, S=2.4 mg/L, SO=2.0 mg/L, and natural infections average S=2.0 mg/L, SO = 1.8 mg/L. A preliminary epitope degradation screen was performed for a panel of antibodies raised to the S1 and S2 regions of the original S protein. The panel showed significant degradation to antibody epitopes in the S1 region. Differential antibody binding of the vaccine response to S and SO suggests vaccine efficacy may be reduced by up to 50% against the Omicron variant.

## Introduction

SARS-CoV-2 has generated several variants of concern^1^, including the Omicron variant (BA.1) with 37 significant mutations to the Spike glycoprotein (SO); 12 of these have been observed in other variants but 25 are unique.^2^ The efficacy of the vaccines to the Omicron variant, or subsequent variant escapes, are then questionable, as the vaccines were designed against the Wuhan variant. National vaccination programmes have now entered ‘booster’ phases to combat waning immunity and improve the protection against Omicron. The global vaccine strategy breaks the link between infection and hospitalisation but is dependent on vaccine efficacy and waning protection. In many countries, population immunity appears to be waning^3^, with vaccine-induced antibody and T cell half-lives reported at a median of 200 days^4^ and at 60 – 200 days for natural antibodies following infection^5^. This range implies immunity waning is clearly a personal endotype characteristic. Consequently, there is a high infection prevalence in the community indicating that any booster programme, unless personalised, will be delivered against a wide spectrum of vaccine and natural infection induced immunity.

Recently, we proposed an antibody mucosal immunity threshold that may be protective against infection based on the concentration of antibodies in the mucosa preventing entrance of the viral particles into the epithelium cells^6^. Concentrations of neutralising and opsonising antibodies in the mucosa may be more than 10^11^ above every mm^-2^ derived from a serum concentration of 1.8 mg/L (95% confidence intervals of 0.2 mg/L – 3.4 mg/L). Taking the upper confidence limit, this predicts an immunity threshold of 3.4 mg/L. The 37 mutations to SO protein may affect the efficacy of antibodies in the mucosa that can bind to the virus and prevent infection, suggesting a higher immunity threshold.

In this paper, we profile the change in epitopes between S and SO proteins for a set of antibodies raised against the original SARS-CoV-2 S glycoprotein to both the S1 and S2 regions of the protein. The assay produces fully quantitative measurements, standardised against the NIST standard human antibody.^7, 8^ We extended the binding study to measure the concentration of antibody binding from three patient cohorts: those who have developed natural immunity following recovery from Wuhan-variant infection early in the pandemic, double-vaccinated patients, and boosted patients. We derive the experimental distribution functions for these three cohorts, review the immunity threshold and consider the evolution of the distribution as antibody levels wane. The differential binding of a patient response to S and SO is considered as a possible predictive measure of vaccine efficacy.

## Methods and Materials

### Methods

#### Biophotonic Multiplexed Immuno-kinetic assay

The immuno-kinetic assay uses a biophotonic detection event and has been described in detail elsewhere in a SARS-CoV-2 antibody sensing application^6, 9^. Briefly, light excites a plasmon wave in the free electrons of gold nanoparticles, which then scatter light. The intensity of scattered light depends on the relative permittivity of the medium in the plasmon field which contains the protein assay; a greater mass of proteins in the plasmon field leads to greater intensity in scattered of light which is measured in real-time. Fundamentally therefore, the biophotonic sensor is a mass sensor calibrated with a monoclonal antibody to a single epitope. The multiplexed nanoparticle array is functionalised with capture molecules to give analytic specificity to the target analyte. The array of the Attomarker COVID-19

Antibody Immunity Test consists of five tests augmented with Protein A/G (PAG) to measure total IgG, polyclonal goat antibodies to measure C-reactive protein (CRP), and the SARS-CoV-2 proteins Nucleocapsid (N), Spike (S), Receptor Binding Domain (RBD) and Omicron Spike (SO) to measure their respective antibodies. In addition, a human serum albumin (HSA) channel acts as a control to adjust for temperature of the array, non-specific binding, and light source intensity variation.

The response of the biophotonic array to S, SO and N is calibrated with 2-point or 3-point calibration using humanised monoclonal antibodies, mass-standardised against the NISTmAb on the PAG sensor binding specifically to the Fc region of the calibration and standard antibodies. NISTmAb is a recombinant humanized IgG1ĸ with a known sequence^10^ specific to the respiratory syncytial virus protein F (RSVF)^8^. RBD is calibrated by comparing the binding site density on the S channel assay with RBD, as determined by the surface coverage of specific non-human antibodies. The analytical specificity of the S and SO proteins was compared using a panel of 7 antibodies with manufacturer defined specificity for S, S1, S2 and RBD, either monoclonal or polyclonal. A further panel of 3 antibodies from SinoBiological was later measured, again against the NIST mAb.

The mucosal immunity threshold^6^ is set at 3.4 mg/L and was derived from experimental distribution function describing the antibody responses of double-vaccinated patients (AZ n = 35, Pfizer n = 25). The reported vaccine efficacies at stopping infection were AZ 69% and Pfizer 93%. Further immunity thresholds were determined using the separation between positive and negative NIBSC samples and the clinical threshold for the Attomarker Triple Antibody Test.

### Materials

Materials were used as supplied by the manufacturer, without further purification. Sigma-Aldrich supplied phosphate buffered saline (PBS) in tablet form (P4417), phosphoric acid solution (85 ± 1 wt. % in water, 345245) and Tween 20 (P1379). Glycine (analytical grade, G/0800/48) was provided by Fisher Scientific. Assay running and dilution buffer was PBS with 0.005 v/v % Tween 20 and the regeneration buffer was 0.1M phosphoric acid with 0.02M glycine solution in deionized water.

The recombinant Human Antibody to the Spike protein S2 subdomain was a chimeric monoclonal antibody (SinoBiological, 40590-D001, Lot HA14AP2901). The antibody was raised against the following immunogen: recombinant SARS-CoV-2 / 2019-nCoV Spike S2 ECD protein (SinoBiological, 40590-V08B). C-reactive-protein-depleted serum was from BBI solutions (SF100-2). NISTmAb was from National Institute of Standards and Technology (RM8671). The detection mixture consisted of a 200-fold dilution of IG8044 R2 from Randox in assay running buffer.

The analytical specificity antibody screening panel was: S2 monoclonal antibody (40590-D001, SinoBiological, lot number HA14AP2901); RBD, S1 monoclonal antibody (40150-D001, SinoBiological, lot number MA14JU0901); S1 monoclonal antibody (40150-R007, SinoBiological, lot number HA14AP3001-B); RBD, S1 and S polyclonal antibody (40589-T62, SinoBiological, lot number HD14AP0705); Polyclonal S2 and S polyclonal antibody (40590-T62, SinoBiological, lot number HD15JU0103); RBD, S1 and S polyclonal antibody (40591-T62, SinoBiological, lot number HD14JU1612); and RBD (Wuhan, Alpha, Beta and Gamma) monoclonal antibody MAB12422 (CR3022, Native Antigen, 21032613). The later panel of three antibodies were all RBD, S1 monoclonal antibodies: (40150-D002, SinoBiological, lot number MA14AP0703); (40150-D003, SinoBiological, lot number HA14AP2304); (40150-D004, SinoBiological, lot number MA14AP0203-B).

Antibody Immunity Test sensor chips were printed with recombinant human serum albumin from Sigma-Aldrich (A9731), anti-CRP from Biorad (1707-0189G), recombinant Protein A/G from Thermo Scientific (21186), SARS-CoV-2 Spike Protein (RBD, His tag) from GenScript (Z03479-100), SARS-CoV-2 Spike S1+S2 ECD-His Recombinant protein from SinoBiological (40589-V08B1), SARS-CoV-2 B.1.1.529 (Omicron) S1+S2 trimer Protein ECD-His from SinoBiological (40589-V08H26), SARS-CoV-2 Nucleocapsid protein from the Native Antigen Company (REC31851-100).

### Patient Samples

#### Commercial samples

Commercial samples purchased from two suppliers (Biomex GmbH and AbBaltis) have previously been used to determine the clinical threshold for the Triple Antibody Test.^6^ A small subset of these were used to assess the antibody response profile following natural infection. All samples tested were found negative for STS, HBsAg, HIV1 Ag (or HIV PCR(NAT)), HIV1/2 antibody, HCV antibody and HCV PCR(NAT) by FDA approved tests; all samples were from RT-PCR positive (PCR+) individuals: 9 samples were purchased from AbBaltis (44% from male donors and 56% from female donors) and 23 were purchased from Biomex (65% from male donors and 35% from female donors).

#### Attomarker Clinic samples

Samples were collected from patients in Attomarker clinics, all of whom have provided informed consent for their anonymised data to be used in research to aid the pandemic response. The data from tests of 48 patient samples are included in this study. 24 had received two doses of either the Oxford-AstraZeneca SARS-CoV-2 vaccine (AZ) or the Pfizer-BioNTech SARS-CoV-2 vaccine (Pfizer/Pf) at least 14 days prior to sample collection and testing by Attomarker. A further 24 had received a third (‘booster’) vaccination of either Pfizer SARS-CoV-2 or Moderna SARS-CoV-2 (Moderna/Mod). The overall vaccine combinations the boosted individuals had received were AZ-AZ-Pf, Pf-Pf-Pf, AZ-AZ-Mod and Pf-Pf-Mod.

## Ethics

The samples were collected with informed patient consent for use in better understanding pandemic. The use of the samples has been reviewed independently by the Biosciences Ethics Committee, University of Exeter and approved.

## Results

An epitope variation profile was obtained by comparing the binding of several commercial antibodies raised to the Wuhan S protein with binding to the Omicron S (calibrated responses). The anti-RBD monoclonal antibody panel binding variations are shown in Table 3, for the anti-RBD antibodies (40150-R007, CR3022, 40150-D002, 40150-D003, 40150-D004) as well as polyclonal anti-S1 antibody, polyclonal anti-S polyclonal and monoclonal anti-S2. The binding ratio shows a ratio range from 1-119, an average of 4.7-fold binding degradation in the RBD neutralising antibody region. The integrity of the mono calibration epitope on the S2 region has degraded, with an estimated affinity of 0.36 nM and 8.0 nM to the SO variant. The integrity of the 3-pt calibration for quantification is maintained^6^ for both the S and SO assays using the same antibody.

**Table 1.** Demographic data for double-vaccinated samples collected in the Attomarker clinic

| Vaccine<br>(doses 1+2) | Total<br>number of<br>donors | Percentage<br>Male | Percentage<br>Female | Number<br>of doses | Min. days<br>since<br>second<br>dose | Max. days<br>since<br>second<br>dose | Mean days<br>since<br>second<br>dose |
| --- | --- | --- | --- | --- | --- | --- | --- |
| ChAdOx1-S<br>(AZ) | 9 | 44.44 | 55.55 | 2 | 18 | 221 | 83 |
| BNT162b2<br>(Pfizer) | 15 | 40 | 60 | 2 | 30 | 224 | 98 |

**Table 2.** Demographic data for booster samples collected in the Attomarker clinic

| Vaccine (dose 3) | Total number of donors | Percentage Male | Percentage Female | Number of doses | Min. days since booster | Max days since booster | Mean days since booster |
| --- | --- | --- | --- | --- | --- | --- | --- |
| BNT162b2 (Pfizer) | 16 | 50 | 50 | 3 | 12 | 115 | 53 |
| Moderna (mRNA-1273) | 8 | 50 | 50 | 3 | 7 | 27 | 21 |

**Table 3.** Differential Epitope Binding Profile

| Spike Region | Anti-RBD |  |  |  |  | Anti-S1 | Anti-S | Anti-S2 |
| --- | --- | --- | --- | --- | --- | --- | --- | --- |
| Antibody | 40150-R007 | CR3022 | 40150-D002 | 40150-D003 | 40150-D004 | Polyclonal | Polyclonal | Monoclonal |
| S/SO Binding | 3.6 ± 0.6 | 10.1 ± 3.2 | 4.2 ± 0.2 | 2.9 ± 0.2 | 2.7 ± 0.2 | 119 ± 1 | 28 ± 5 | 1.9 ± 0.1 |

The antibody response distributions from the three patient cohorts for both the S and SO proteins are shown in Figure 1, demonstrating that the booster response ranges from 0.5 mg/L – 100 mg/L for the S protein and 0.5 mg/L – 140 mg/L for the SO protein. The averages (mode) of the booster cohort distributions were 15.1 mg/L and 13.4 mg/L, 89% lower for S and SO, respectively, compared with the significantly lower double-vaccinated mode, S = 2.4 mg/L, SO = 2.0 mg/L and natural infection mode S = 2.0 mg/L and (SO) = 1.8 mg/L.

**Figure 1.**
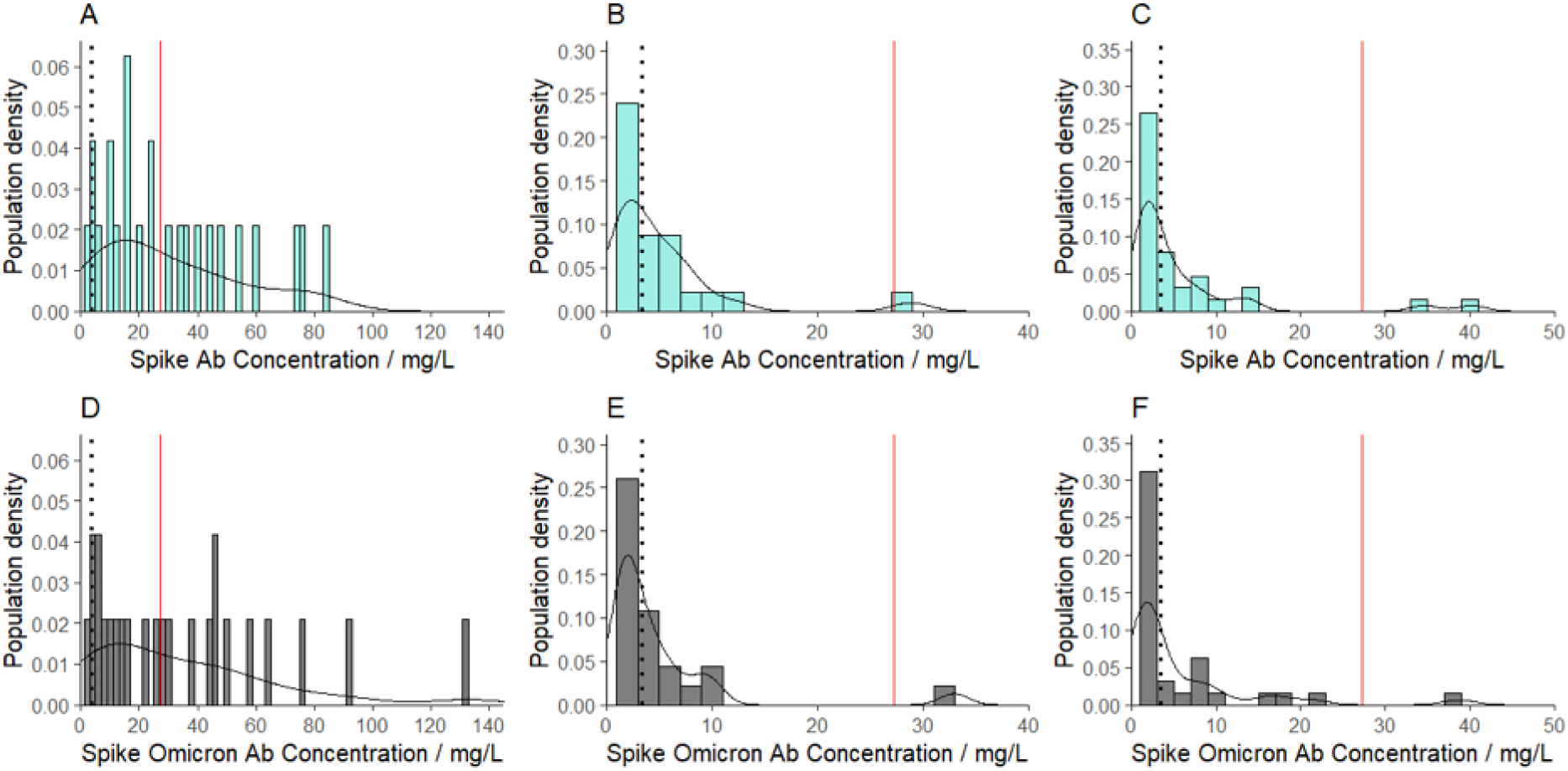
The fully quantitative experimental response distribution functions for: (A) booster response to S protein; (B) double-vaccinated response to S protein; (C) natural immunity response to S protein; (D) booster response to SO protein; (E) double-vaccinated response to SO protein; and (F) natural immunity response to SO protein. The dotted line is the immunity threshold at 3.4 mg/L and the solid line is set at 27.2 mg/L: 8-times (or 3-half-life periods) above the immunity threshold.

Among the experimental vaccine booster responses in all dose regimen combinations, 97.5% are above the mucosal immunity threshold for S and 95.1 % for SO. The same figures for the double-vaccinated cohort return 51.9 % for S and 44.3 % for SO. Finally, the patients recovering from a natural infection return 44.6 % above the mucosal threshold for S and 35.7 % for SO.

The ratio of binding (SO/S) is also shown by vaccine type for the booster and double-vaccinated cohorts and for the natural immunity cohort, Figure 2. There is no consistent variation between vaccine sequence but there are larger differences in the ratio range with a maximum distribution range over an order of magnitude. The double-vaccinated and natural immunity cohorts include more patients with the SO/S ratio below 0.5.

**Figure 2.**
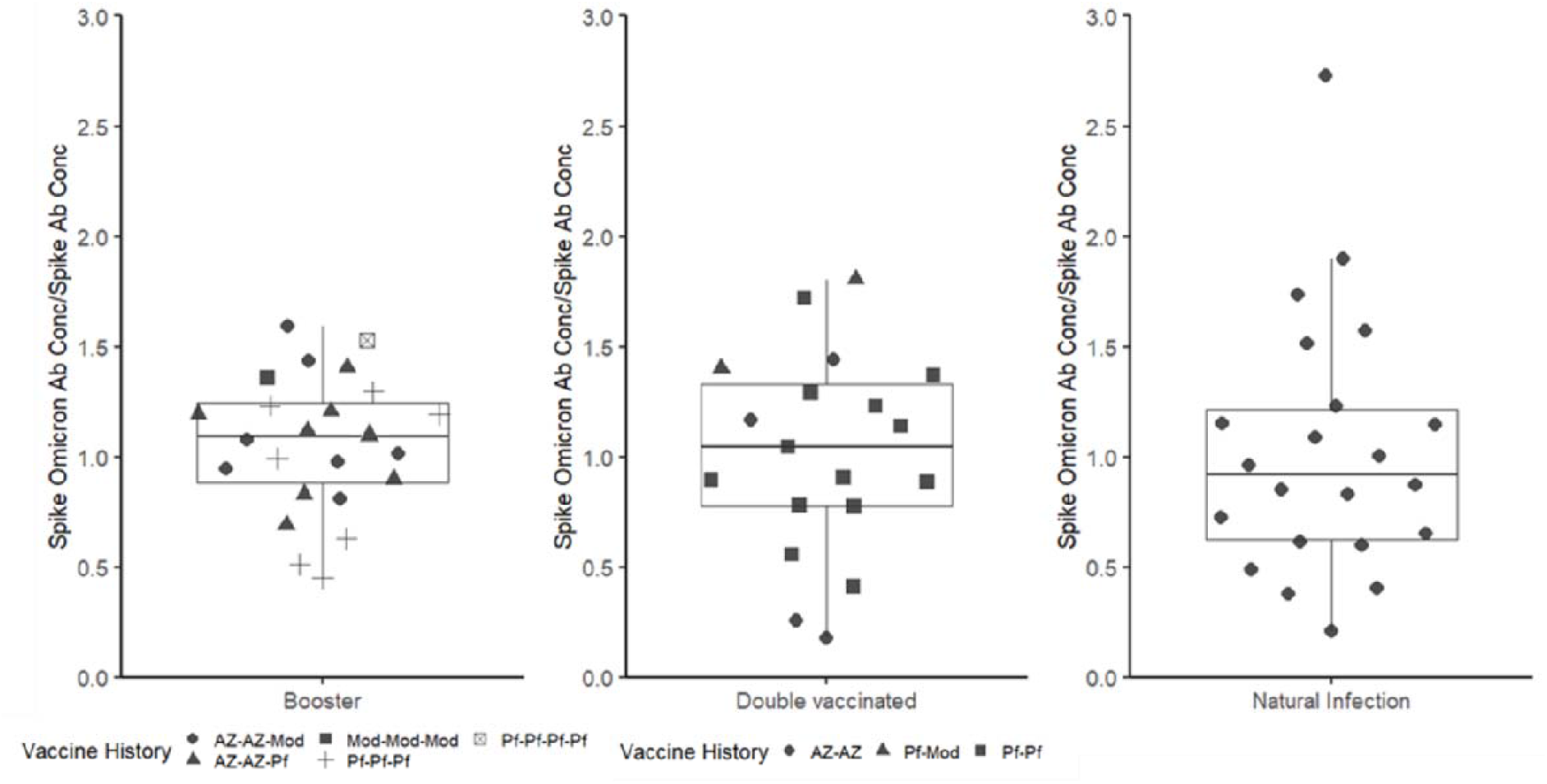
Bee-swarm boxplot for the ratio SO/S for the three patient cohorts, showing the different vaccination regimens.

## Discussion

The SO protein has 37 significant mutations: 12 of these have been observed in other variants of concern, but 25 are unique. Antibody epitopes are a function of 4-or 6-amino acid sequences forming a linear epitope, modified by the extensive glycosylation patterns; formally however, epitopes are electrostatic patches on the surface of the target protein. Degradation of the antibody epitopes from their original sites on the S protein to the variant SO protein was screened herein using a set of antibodies raised to the S protein from the Wuhan variant, some polyclonal and some monoclonal, including the calibration antibody. The calibration antibody to the S2 region showed a decrease in affinity by 40%: from 2.7 ± 0.30 nM to 1.3 ± 0.13 nM. The S1 region of the SO protein shows significant binding differences across the antibody panel, most pronounced for the RBD region which shows a 119-fold reduction in binding for one antibody. The RBD region is the target of neutralising antibodies specifically binding with the ACE2 receptor, the entry point by which SARS-CoV-2 targets epithelial cells.^11, 12, 13^ The vaccine efficacy as determined by the ratio of SO/S binding varies between 0.5 and 1.5 in the booster population suggesting a proportion of the population with vaccinated immunity will have degraded vaccine efficacy against the omicron variant. Further, given the degradation to RBD, the booster population has a significant proportion of affinity-matured antibodies that are not directly bound to the RBD region but are still potentially protective sterically. Opsonisation is also a likely protective mechanism in the mucosa, causing recruitment of Complement proteins to antibody complexes.

The concentrations of antibodies produced by the double-vaccinated and convalescent cohorts are significantly lower for S and SO than responses to the booster. Importantly, samples from the double-vaccinated cohort were taken approximately two weeks after the second vaccination and represent peak immunity. For the booster cohort however, the third dose was given to all patients regardless of their immunity status at the time. In part, the unknown immunity status of the boosted patients pre-third dose explains the large range in their responses: from 0.5 mg/L – 100 mg/L for the S protein and 0.5 – 140 mg/L for the SO protein. This range is greater for SO than for S, although the sample size is small. All but the lower 5% of the booster patient responses are above our proposed mucosal Immunity Threshold, suggesting protection from infection. The spectrum of response is likely the product of boosting individuals already on a wide spectrum of pre-booster immunity, as well as the different combinations of vaccines received, e.g., AZ-AZ-Pf, Pf-Pf-Pf, AZ-AZ-Mod and Pf-Pf-Mod. Prior to boosting, 40% of patients attending Attomarker clinics had responses below the threshold, with anecdotal reporting of RT-PCR-confirmed cases for COVID-19. In addition to the combinations of vaccination, increases in antibody levels from primary natural infection and re-exposure will contribute to a diverse immunity background and antibody response.

Antibody levels as correlates of immunity to SARS-CoV-2 have been studied extensively^14,15^, as well as for 18 previous vaccines^16, 17^. Vaccine efficacy against Omicron and how long immunity will last are key questions for management of the pandemic. First-generation vaccines raised against the S protein had impressive efficacies even at the lower antibody concentrations, Figure 2 (B) and (E); the Pfizer vaccine has an efficacy of 93% for reducing symptomatic cases^18^ ≥14 days following the second dose; AZ with an equivalent efficacy of 69% – 74% in a real-world setting^19^; and Moderna showing 95.2% efficacy (95% confidence limits 91.2 % – 97.4 %)^20^. This immunity, consisting of antibodies and T cell responses, wanes over time.

The booster response is complex and diverse across individuals^21^, with differently reported antibody half-lives. The half-life for IgG antibodies following SARS-CoV-2 infection varies between 60 – 200 days, with a mean of 106 days^5^. A further study estimated the half-life of IgG from the peak Pfizer vaccine-induced humoral response to be 21 (95% CI 13 – 65) days in initially seronegative individuals, but 53 (95% CI 40 – 79) days in initially seropositive individuals. The estimated half-life for total antibodies was longer and ranged from 68 (95% CI 54 – 90) days to 114 (95% CI 87 – 167) days in initially seronegative and initially seropositive individuals, respectively^11^, with a median half-life of 110 days. Thus, protection from the booster may already be significantly lower in patients who received their booster first, including many healthcare professionals and the clinically vulnerable.

The complexity of responses and vaccination histories in the community suggests a need to profile antibody concentrations in individuals, which could provide a useful insight into evolving population immunity. Consider the patient in this study who returned a serum antibody concentration of 13 mg/L: predicting a half-life of 100 days, they would have 200 immunity days before falling below the mucosal immunity threshold. However, if the individual had a half-life of 60 days (the lower quartile), they would need a booster sooner, in approximately 120 days. Alternatively, the Pfizer booster patient with a serum antibody concentration of 37 mg/L (more than 9 times higher than the mode concentration ≥14 days post-first vaccine dose) prompts questions around whether a safe upper antibody limit for further vaccination exists, and the consequent risk of autoimmune diseases^22^.

## Conclusions

The spectrum of vaccine responses does suggest that for some individuals, immunity above the mucosal immunity threshold may last for extended periods. A concentration of 27 mg/L, for example, is 8-times the mucosal immunity threshold and antibodies would remain above it for 3-half-lives or 180 days. For the median-half-life patient this would equate to approximately one year – their ‘safe zone’. The vaccine efficacy against the omicron variant will be lower for some patients suggesting a higher mucosal immunity threshold up to 7.4 mg/L if the efficacy is reduced by 50%, as indicated by the data, and a consequently shorter period of protection from symptomatic infection. By extension, vaccine escape could be defined as a higher mucosal threshold from a new variant, which would leave more than 50% of the population without sufficient antibodies.

Personalised immunity profiling with quantitative antibody testing is possible on a population level and could be used to assess the need for subsequent boosters. It is also important to consider whether patients with high antibody levels, such as 140 mg/L, and long half-lives of 200 days or more may be at risk from over-boosting or ‘overdosing’, with potential risks of autoimmune diseases. Therefore, personal, precision immunity profiling may form a part of ongoing endemic management.

## Data Availability

All data produced in the present study are available upon reasonable request to the authors

## Acknowledgements

The authors would like to thank Dr Jonathan Snicker and Mr Sydney Nash for their guidance on the potential strategic and policy implications of the scientific findings.

## References

1. Hayawi, K., Shahriar, S., Serhani, M.A., Alashwal, H. & Masud, M.M. Vaccine versus Variants (3Vs): Are the COVID-19 Vaccines Effective against the Variants? A Systematic Review. Vaccines 9 (2021).

2. Sarkar, R., Lo, M., Saha, R., Dutta, S. & Chawla-Sarkar, M. S glycoprotein diversity of the Omicron Variant. medRxiv, 2021.2012.2004.21267284 (2021).

3. Goldberg, Y. et al. Waning Immunity after the BNT162b2 Vaccine in Israel. The New England journal of medicine (2021).

4. Cohen, K.W. et al. Longitudinal analysis shows durable and broad immune memory after SARS-CoV-2 infection with persisting antibody responses and memory B and T cells. Cell Rep Med 2, 100354–100354 (2021).

5. Dan, J.M. et al. Immunological memory to SARS-CoV-2 assessed for up to 8 months after infection. Science 371, eabf4063 (2021).

6. James-Pemberton, P.H. et al. Fully Quantitative Measurements of the Antibody Levels for SARS-CoV-2 Infections and Vaccinations calibrated against the NISTmAb Standard IgG Antibody – an Immunity Test Threshold. New England Journal of Medicine (Submitted) (2021).

7. Schiel, J.E. et al. The NISTmAb Reference Material 8671 value assignment, homogeneity, and stability. Analytical and bioanalytical chemistry 410, 2127–2139 (2018).

8. McLellan, J.S. et al. Structural basis of respiratory syncytial virus neutralization by motavizumab. Nature structural & molecular biology 17, 248–250 (2010).

9. Shaw, A.M. et al. Real-world evaluation of a novel technology for quantitative simultaneous antibody detection against multiple SARS-CoV-2 antigens in a cohort of patients presenting with COVID-19 syndrome. Analyst (2020).

10. Formolo T L.M., Levy M, Kilpatrick L, Lute S, Phinney K, et al. Determination of the NISTmAb primary structure. State-of-the-Art and Emerging Technologies for Therapeutic Monoclonal Antibody Characterization, vol. 2. ACS Symposium Series, 2015, pp 1–62.

11. Bayart, J.L. et al. Waning of IgG, Total and Neutralizing Antibodies 6 Months Post-Vaccination with BNT162b2 in Healthcare Workers. Vaccines 9 (2021).

12. Khoury, D.S. et al. What level of neutralising antibody protects from COVID-19? medRxiv, 2021.2003.2009.21252641 (2021).

13. Nguyen, D. et al. SARS-CoV-2 neutralising antibody testing in Europe: towards harmonisation of neutralising antibody titres for better use of convalescent plasma and comparability of trial data. Euro surveillance : bulletin Europeen sur les maladies transmissibles = European communicable disease bulletin 26 (2021).

14. Abbasi, J. The Flawed Science of Antibody Testing for SARS-CoV-2 Immunity. Jama 326, 1781–1782 (2021).

15. Earle, K.A. et al. Evidence for antibody as a protective correlate for COVID-19 vaccines. Vaccine 39, 4423–4428 (2021).

16. Thakur, A., Pedersen, L.E. & Jungersen, G. Immune markers and correlates of protection for vaccine induced immune responses. Vaccine 30, 4907–4920 (2012).

17. Plotkin, S.A. Correlates of protection induced by vaccination. Clin Vaccine Immunol 17, 1055–1065 (2010).

18. Dagan, N. et al. BNT162b2 mRNA Covid-19 Vaccine in a Nationwide Mass Vaccination Setting. New England Journal of Medicine 384, 1412–1423 (2021).

19. Mallapaty, S. & Callaway, E. What Scientists do and don’t know about the Oxford-AstraZeneca COVID vaccine. Nature 592, 15–17 (2021).

20. Baden, L.R. et al. Efficacy and Safety of the mRNA-1273 SARS-CoV-2 Vaccine. The New England journal of medicine 384, 403–416 (2021).

21. Barrett, J.R. et al. Phase 1/2 trial of SARS-CoV-2 vaccine ChAdOx1 nCoV-19 with a booster dose induces multifunctional antibody responses. Nat Med 27, 279–288 (2021).

22. Dotan, A. et al. The SARS-CoV-2 as an instrumental trigger of autoimmunity. Autoimmunity reviews 20, 102792 (2021).

